# Multimodal Speech Biomarkers for Remote Monitoring of ALS Disease Progression

**DOI:** 10.1101/2024.06.26.24308811

**Authors:** Michael Neumann, Hardik Kothare, Vikram Ramanarayanan

**Affiliations:** Modality.AI, Inc., San Francisco, CA, USA; University of California, San Francisco, CA, USA

**Author notes:** These authors contributed equally to the work (Michael Neumann, Hardik Kothare).

**Keywords:** Remote Patient Monitoring, Multimodal Digital Biomarkers, Explainability, Biomedical Speech, Voice Signal Processing

## Abstract

Amyotrophic lateral sclerosis (ALS) is a progressive neurodegenerative disease that severely impacts affected persons’ speech and motor functions, yet early detection and tracking of disease progression remain challenging. The current gold standard for monitoring ALS progression, the ALS functional rating scale - revised (ALSFRS-R), is based on subjective ratings of symptom severity, and may not capture subtle but clinically meaningful changes due to a lack of granularity. Multimodal speech measures which can be automatically collected from patients in a remote fashion allow us to bridge this gap because they are continuous-valued and therefore, potentially more granular at capturing disease progression. Here we investigate the responsiveness and sensitivity of multimodal speech measures in persons with ALS (pALS) collected via a remote patient monitoring platform in an effort to quantify how long it takes to detect a clinically-meaningful change associated with disease progression. We recorded audio and video from 278 participants and automatically extracted multimodal speech biomarkers (acoustic, orofacial, linguistic) from the data. We find that the timing alignment of pALS speech relative to a canonical elicitation of the same prompt and the number of words used to describe a picture are the most responsive measures at detecting such change in both pALS with bulbar (*n* = 36) and non-bulbar onset (*n* = 107). Interestingly, the responsiveness of these measures is stable even at small sample sizes. We further found that certain speech measures are sensitive enough to track bulbar decline even when there is no patient-reported clinical change, i.e. the ALSFRS-R speech score remains unchanged at 3 out of a total possible score of 4. The findings of this study have the potential to facilitate improved, accelerated and cost-effective clinical trials and care.

## 1. ALS & Speech Biomarkers

Amyotrophic Lateral Sclerosis (ALS) is a progressive motor neuron disease with an estimated global prevalence of 4.42 per 100,000 persons [1]. Neuronal death leads to muscular atrophy, loss of voluntary motor control in persons with ALS (pALS) and a median survival of 3 to 5 years [2] after disease onset. Up to 30% of pALS present with bulbar onset of ALS, characterised by a rapid loss of speech and swallowing functions [3], while the rest present with non-bulbar onset characterised by muscular atrophy in the limbs and the trunk [4]. However, a vast majority of non-bulbar onset pALS eventually also exhibit bulbar symptoms in the course of their disease progression [2]. The heterogeneous nature of ALS onset and progression underlines the importance of identifying efficacious biomarkers to improve the predictive modelling of disease progression.

The current clinical gold standard to track disease progression in ALS is the ALS Functional Rating Scale - Revised (ALSFRS-R) [5], a questionnaire comprising 12 questions across four functional domains impacted by ALS [6]: bulbar, fine motor, gross motor and respiratory. However, there is evidence that the ALSFRS-R scale may track disease progression in a non-linear manner and may lack sensitivity in the early stages of bulbar disease onset [7, 8, 9]. For example, Van Unnik et al. have pointed out that such survey-based outcomes have “limited ability to detect subtle changes over time” [10].

Speech and oro-facial biomarkers have shown great promise for remote assessment and monitoring of neurological and mental health [11, 12, 13, 14, 15]. Indeed, many studies have computed and demonstrated the efficacy of multiple speech metrics that capture how a given disease impacts multiple domains of speech performance – be it motor, anatomical, cognitive, linguistic or affective [11, 16, 17, 18]. Objective speech and facial kinematic measures have been shown to be very powerful in early detection of bulbar symptoms [19, 20, 21, 22, 23, 24, 25] and the progression of bulbar decline in pALS [26, 27, 28, 29, 30]. Eshghi et al. [29] demonstrated that speaking rate and speech intelligibility can predict speech loss based on pre-defined thresholds and that these objective speech measures are more responsive to functional decline than patient-reported ALSFRS-R scores. Yunusova et al. [27] suggested that changes in kinematics of the jaw and lips are detectable prior to changes in vowel acoustics and speech intelligibility. Stegmann et al. [28] demonstrated that disease progression in bulbar onset and non-bulbar onset pALS can be predicted using speaking rate and articulatory precision through data collected remotely via a mobile application. Speaking rate has been consistently found to be an important biomarker for early diagnosis and stratification in both these studies and other studies, along with other timing-related measures like percentage pause time, speaking duration and others [22, 23, 31, 32]. Prior work by us [30] has shown that some timing-related speech biomarkers, collected remotely through a conversational dialog platform, have the requisite responsiveness and sensitivity to track speech decline in the context of clinical interventional trials targeting neurodegenerative disorders. To establish the efficacy of multimodal biomarkers in tracking disease progression, it is important to consider what constitutes a minimal clinically-important difference (MCID) [33, 34, 35] and whether these biomarkers show change greater than any measurement errors. It is important that these multimodal biomarkers are also sensitive in detecting bulbar decline, which could be well before corresponding changes are observed in the relevant ALSFRS-R functional scores or equivalent clinical scales.

To address the need for improved biomarkers of bulbar disease progression in ALS, we explored the responsiveness, sensitivity and clinical utility of multimodal speech metrics^1^ – automatically extracted via a cloud-based multimodal dialog platform – by formulating the following research questions:

1. *Optimal Feature Selection*. Which speech and facial biomarkers are the best at capturing differences between bulbar and non-bulbar onset ALS?
2. *Responsiveness to Longitudinal Change*. How is the rate of change in these speech and facial biomarkers different for bulbar and non-bulbar onset pALS? Can we quantify how different the rates are?
3. *Time to Detect Change*. How many weeks does it take to detect a clinically meaningful change in these biomarkers from disease onset in both cohorts of pALS, keeping in mind that ALS is a rapidly-progressing disease?
4. *Effect of Sample Size*. How does the responsiveness and time to detect change depend on the sample size of the cohort?
5. *Sensitivity Relative to Clinical Standard*. Can these metrics detect speech deterioration during intervals of time when patients report no speech changes on the ALSFRS-R instrument?

The present study is partially inspired by the work by Stegmann et al. [28] who applied growth curve models to model longitudinal trajectories of speaking rate and articulatory precision in pALS with bulbar onset and non-bulbar onset. We extend this work in several key aspects: (i) by analyzing a considerably larger dataset (278 participants total) that was recorded using an interactive dialog system; (ii) by exploring a large multimodal feature set to find the most promising features for the task at hand; (iii) by conducting a sample size analysis; and (iv) by putting the longitudinal modelling results into context with respect to MCID thresholds (based on work by Stipancic et al. [34]) and ALSFRS-R scores. As mentioned previously, we have addressed some of these research questions using a very narrow set of speech timing features in [30]. However, this is the first data-driven study, to the best of our knowledge, to look at the responsiveness and sensitivity of remotely-collected multimodal (i.e., speech, facial and text based) digital biomarkers extracted using a structured conversational dialog with a virtual agent. The advantage of such self-driven assessments is that there is no software or hardware installation required and data collection can be done using the participants’ devices.

The rest of this paper is organized as follows. Section 2 provides information about the study design and the dataset. The dialog system that was used for data capture is described in section 3. Section 4 presents all methodological details, including automatic feature extraction, feature selection, MCID estimation, and the longitudinal analysis based on growth curve models. Our findings are presented in section 5, before we conclude this paper with the discussion in section 6.

## 2. Data and Study Design

The study protocol was approved by an external Institutional Review Board^2^ on August 11, 2020. Participants were recruited by EverythingALS and the Peter Cohen Foundation^3^. All participants provided informed consent upon recruitment, prior to their first assessment on the Modality platform. Figure 1 provides an overview of the study procedures. Data was collected between 2020-11-03 and 2023-10-06 from 143 pALS (70 female, mean age *±* standard deviation = 60.4 *±* 10.2 years, Bulbar onset: *n* = 36, Non-Bulbar onset: *n* = 107) and 135 age and sex-matched controls (71 female, mean age *±* standard deviation = 59.9 *±* 10.3 years). For age matching, a tolerance threshold of *±*3 years was set^4^. A total number of 6,816 recording sessions were conducted (3,388 sessions from pALS – 598 bulbar onset and 2,790 non-bulbar onset – and 3,428 sessions from healthy controls). Out of 3,388 sessions from pALS, the ALSFRS-R total score was available for 1,879 sessions. Table 1 summarizes the participant statistics and provides information on the ALSFRS-R scores at baseline (participants’ first session).

**Figure 1:**
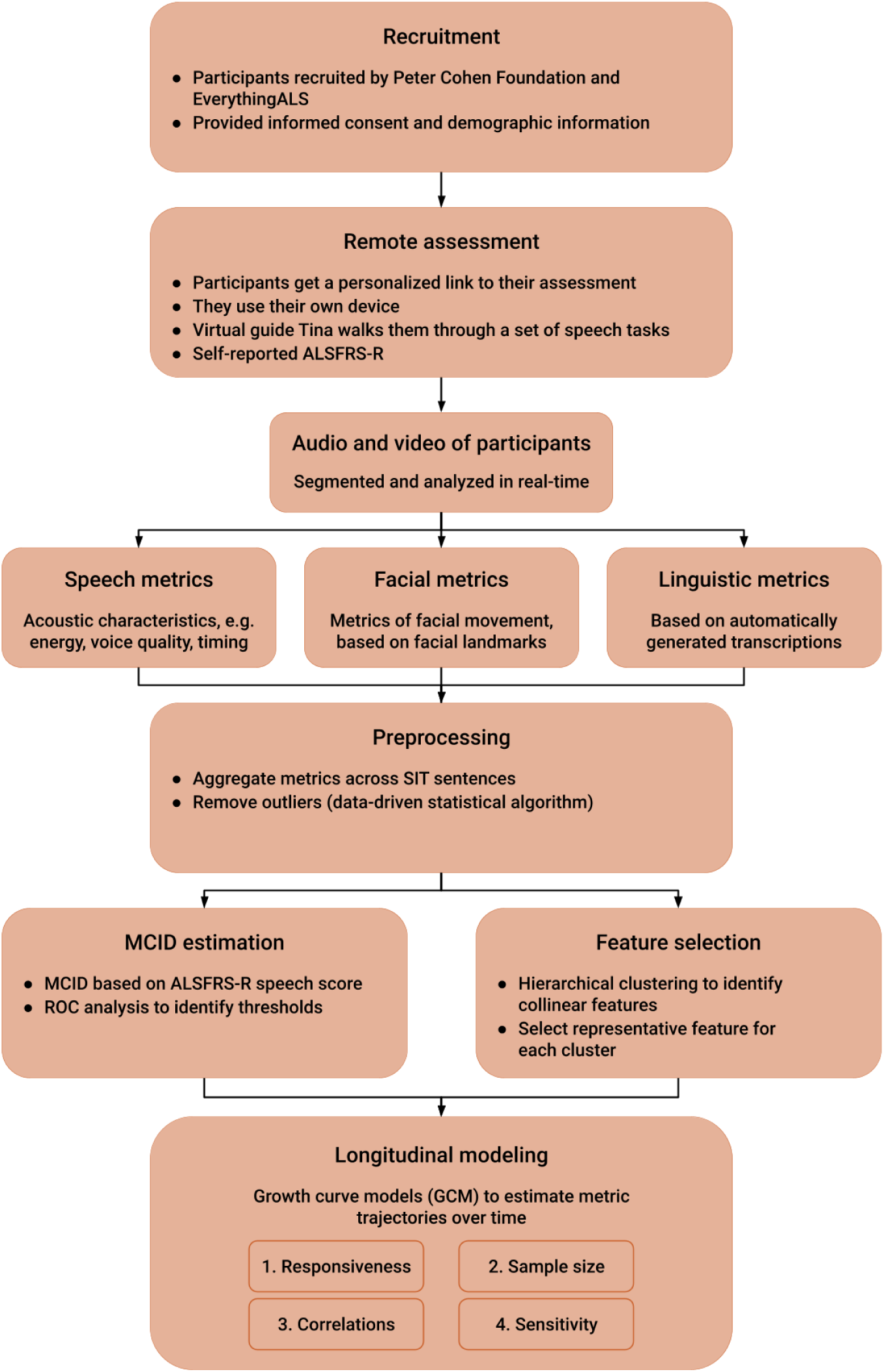
Schematic of study design and modeling procedures.

**Table 1:** Participant statistics. F: number of female participants, M: number of male participants, BL: baseline (first session). The time span *first-to-last* is the mean number of days between participants’ first and last session in the data collection. The large variation of the number of samples per participant in this data collection is due to the continuous and ongoing recruitment of new participants in this study. Statistics are reported as *median; mean (standard deviation)* for ALSFRS-R scores, and as *mean (standard deviation)* otherwise.

|  | F | M | Age (years) | #Sess. | ALSFRS-R<br>Baseline | ALSFRS-R<br>speech BL | #Sessions<br>per participant | Time span<br>first-to-last (days) |
| --- | --- | --- | --- | --- | --- | --- | --- | --- |
| Bulbar onset | 18 | 18 | 61.6 (11.9) | 598 | 38.5; 38.9 (6.1) | 3.0; 2.7 (0.8) | 16.6 (19.4) | 232.8 (222.2) |
| Non-bulbar onset | 52 | 55 | 59.9 (9.6) | 2,790 | 38.0; 35.6 (7.4) | 4.0; 3.6 (0.6) | 26.1 (25.9) | 377.5 (305.1) |
| All pALS | 70 | 73 | 60.4 (10.2) | 3,388 | 38.0; 36.4 (7.2) | 4.0; 3.4 (0.8) | 23.7 (24.7) | 341.1 (292.6) |
| Healthy controls | 71 | 64 | 59.9 (10.3) | 3,428 | - | - | 25.4 (24.5) | 317.7 (262.6) |

## 3. Multimodal Dialog System

The Modality service, a cloud-based multimodal dialog system [36, 37, 38], was used to collect video recordings from participants, who engaged in a structured conversation with Tina, a virtual dialog agent (see Figure 2 for a schematic illustration of the dialog platform). To ensure data privacy and protection of personal health information (PHI), the Modality service is fully compliant with the Health Insurance Portability and Accountability Act (HIPAA) and the General Data Protection Regulation (GDPR; European Union). Each participant is provided with a unique website link to the Modality platform, which they can click on to start the assessment using a browser and device of their choice (microphone and webcam required).

**Figure 2:**
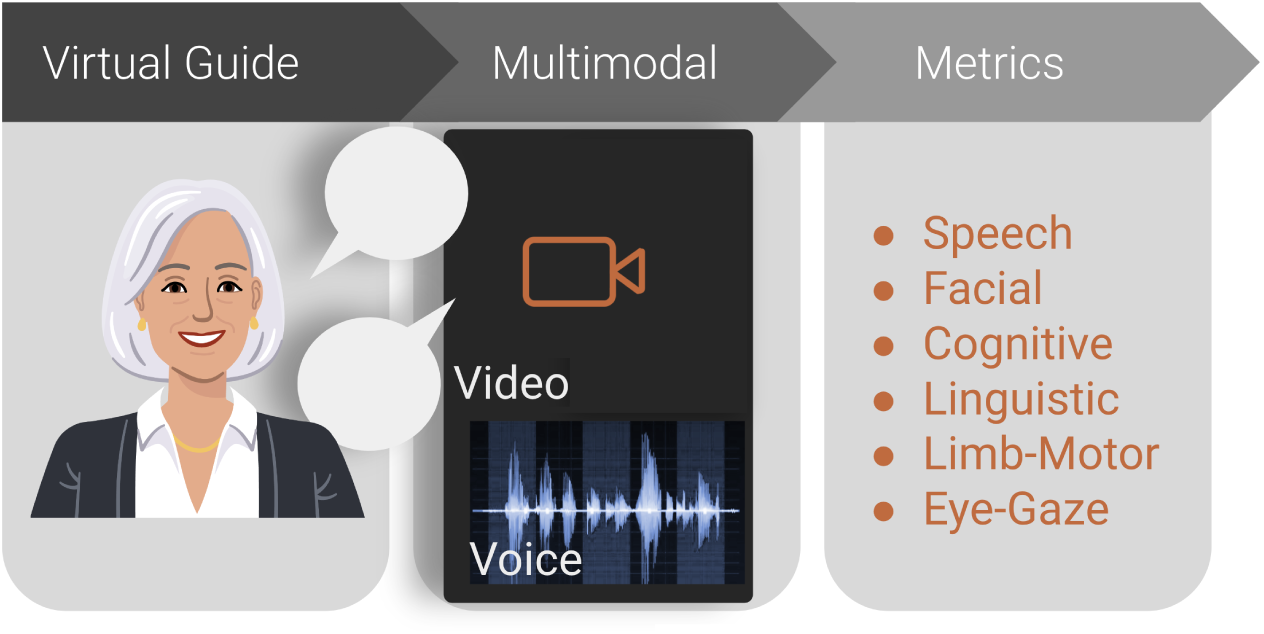
Schematic diagram of the Modality dialog platform.

After completing microphone and camera checks to ensure data collection of good quality, participants engage in a conversation with Tina. The dialog protocol elicits different types of speech samples that are inspired by prior work [39, 40, 41, 42] and also utilized in similar remote monitoring efforts [15, 43]. In this data collection, the following tasks were included: (a) read speech (sentence intelligibility test (SIT), 5-15 words; Bamboo reading passage (RP), 99 words), (b) measure of diadochokinesis (DDK, rapidly repeating the syllables /pataka/), (c) single breath counting (SBC), and (d) free speech in form of a picture description task (PD). During the assessment, Tina asks participants to do the aforementioned tasks. Due to the conversational nature, participants receive feedback (e.g., when they spoke shorter than a predefined threshold for a given task), and Tina can provide demonstrations of how a task should be performed. Participants’ audio and video streams are uploaded to the cloud and segmented in real-time for downstream analysis. After dialog completion, participants were asked to fill out the ALS functional rating scale - revised (ALSFRS-R) [5], the standard clinical scale to capture progression in ALS.^5^

## 4. Methods

### 4.1. General Experimental Setup

All analyses were performed using Python (v3.10) and R (v4.3.1). The following open-source Python libraries were used: Pandas (v1.5.3 [45, 46]), Numpy (v1.24.3 [47, 48]), scikit-learn (v1.2.2 [49]), Matplotlib (v3.7.1 [50]), spaCy^6^ (v3.5.3), and SciPy (v1.10.1 [51]). The following R packages were used: ROCR (v1.0.11 [52]), pROC (v1.18.2 [53]), ggplot2 (v3.4.4 [54]), nlme (v3.1.162 [55]), and the rpy2^7^ interface (v3.5.13).

### 4.2. Speech and Facial Metrics

Our multimodal dialog platform is equipped with analytics modules that automatically extract metrics to capture information from acoustic (energy, timing, voice quality, spectral), facial (articulatory kinematics, range of motion, eye and facial movement), motoric (finger tapping kinematics) and textual (lexico-semantic, sentiment) domains during the different tasks. Table 2 provides an overview of the extracted metrics.

**Table 2:** Overview of extracted metrics. For visual metrics, functionals (minimum, maximum, average) are applied to produce one value across all video frames of an utterance. Visual distance metrics are measured in pixels and are normalized by dividing them by the intercanthal distance (distance between inner corners of the eyes) for each participant. *specific to DDK task

|  | Domain | Exemplar Metrics |
| --- | --- | --- |
| *Audio* | Energy | shimmer (%), intensity (dB), signal-to-noise ratio (dB) |
|  | Timing | speaking and articulation duration (sec.), articulation and speaking rate (WPM), percent pause time (PPT, %), canonical timing agreement (CTA, %), cycle-to-cycle temporal variability* (cTV, sec.), syllable rate* (syl./sec.), number of syllables* |
|  | Voice quality | cepstral peak prominence (CPP, dB), harmonics-to-noise ratio (HNR, dB) |
|  | Frequency | mean, max., min. fundamental frequency F0 (Hz), first three formants F1, F2, F3 (Hz), slope of 2nd formant (Hz/sec.), jitter (%) |
| *Text* | Lexico-semantic | word count, percentage of content words, noun rate, verb rate, pronoun rate, noun-to-verb ratio, noun-to-pronoun ratio, closed class word ratio, idea density |
| *Video* | Mouth (distances) | lip aperture/opening, lip width, mouth surface area, mean symmetry ratio between left and right half of the mouth |
|  | Lip/Jaw Movement | velocity, acceleration, jerk, and speed of lower lip and jaw center |
|  | Eyes | number of eye blinks per sec., eye opening, vertical displacement of eyebrows |

We use Praat (v6.2.17) [56] and the Montreal Forced Aligner (v2.0.0.a22) [57] to extract speech metrics, including timing measures, such as percentage of pause time (PPT; proportion of the total duration of all pauses to the total duration of the utterance), rate measures such as speaking rate (the total number of words in the passage (99) divided by the speaking duration, or time taken to read the Bamboo passage [58]), frequency related measures, such as fundamental frequency (F0), energy related measures, such as signal-to-noise ratio, and voice quality measures, such as the harmonics-to-noise ratio (HNR). We also computed Canonical Timing Alignment (CTA; %), a number between 0% (non-alignment) and 100% (perfect alignment), measured as the normalised inverse Levenstein edit distance between words and silence boundaries (here the participant’s predicted word-level timing information, derived using the Montreal Forced Aligner [59] is compared to a canonical production by Tina [60]).

Facial video metrics are based on facial landmarks generated with MediaPipe Face Mesh [61]. First, MediaPipe Face Detection, which is based on BlazeFace [62], is used to determine the (x, y)-coordinates of the face for every video frame. Then, facial landmarks are extracted using MediaPipe Face Mesh. We use 14 key landmarks to compute metrics like the speed of articulators (jaw, lower lip), surface area of the mouth, and eyebrow raises. These landmarks include center and corners of the lips, jaw center, nose tip, center and corners of the eyes, and the center of the eyebrows. Lastly, the features are normalized by dividing them by the inter-caruncular or inter-canthal distance, to handle variability across participant sessions due to position and movement relative to the camera [63].

Linguistic metrics are computed for the picture description task only, using the Python package spaCy. They are based on automatic transcriptions obtained with AWS Transcribe^8^ and include lexico-semantic metrics, such as word count, noun rate, and noun-to-verb ratio.

### 4.3. Preprocessing

Generally, every metric is computed for each task of the assessment on an utterance level, e.g., speaking rate for the reading passage, or speaking duration for the SBC task. In the present work, we refer to these task-metric combinations as *features*. For the SIT task, metrics were aggregated across six SIT sentences by taking the mean values over these utterances (i.e., speaking rate for SIT denotes the average speaking rate across the six sentences).

To remove outlier values from speech and facial features, we employed a distribution-based outlier detection algorithm [64]. Possible reasons for outlier occurrence include high-intensity background noise, bad lighting conditions, or incorrectly performed tasks. First, all feature values that are more than five standard deviations away from the population mean are removed. These are considered extreme outliers, which potentially skew the distribution mean. Such extreme events can happen when the recorded data is corrupted, for example through a poor network connection. The value of five standard deviations was empirically chosen after carefully analyzing the data distributions. Then, the mean is re-computed and values outside *±*3 standard deviations are flagged as outliers and removed from any further analysis.

For the feature selection procedure (section 4.5), all features were normalized to zero mean and unit variance. For the longitudinal analysis, where we look at one feature at a time, we decided to work with raw, unnormalized values because this helps with interpreting the intercepts and slopes of the growth curve models.

### 4.4. Clinically-meaningful change

To clearly define what feature changes count as clinically meaningful, we use the concept of the minimal clinically-important difference (MCID) [33, 34]. The MCID is the smallest domain-specific change that is considered to be clinically relevant [65]. It can be quantified as a threshold for a change corresponding to clinical improvement or deterioration [35] and is tied to an external anchor, which is considered to be a clinical gold standard, the ALSFRS-R speech question in this case. We calculated the MCID for all features for a corresponding one-point change on the ALSFRS-R speech question where participants are asked to rate their speech on the following scale with scores in parentheses:

- Normal speech processes (4)
- Detectable speech disturbance (3)
- Intelligible with repeating (2)
- Speech combined with nonvocal communication (1)
- Loss of useful speech (0)

One approach to derive the MCID is using data-driven ROC analysis [66], which was also applied in [34]. The point representing maximum sensitivity and specificity (closest to the top left corner) on an ROC curve is determined as the optimal cutpoint corresponding to the MCID value. MCID calculation was performed using the rpy2 package in Python along with the pROC [67] and ROCR [52] packages in R [68]. The classes being discriminated were pALS who exhibited a one-point decline in their ALSFRS-R speech score and those who did not show any change in their ALSFRS-R speech score. For each pALS, adjacent sessions (with at least 14 days between sessions) were considered to calculate the change in every feature from the first to the second session. For pALS in the one-point decline class, only those adjacent sessions were taken into account where the decline was observed.

### 4.5. Feature Selection

All audiovisual metrics were extracted for each of the five speech tasks in the protocol. Considering all valid task-metric combinations as individual features results in a very large number of features. To handle multicollinear features and identify a good set of representative features, we applied hierarchical clustering on the Spearman rank-order correlations, similar to the approach in [69]. For this feature clustering approach, only healthy controls’ data was considered in order to avoid data leakage in the experimental design – note that all subsequent analyses focuses on patient data only – and because data from healthy controls is most representative of normative feature ranges and correlations between features. Ward’s method was used for clustering and we plotted a dendrogram for visual inspection of the feature clusters (see Figure 3). A distance threshold of 1.0 to split clusters^9^ was chosen manually to select clusters that represent sensible feature groupings in terms of the domain (e.g. frequency or timing related speech features) or the area of the face (e.g. features pertaining to jaw movement). This threshold resulted in 27 clusters.

**Figure 3:**
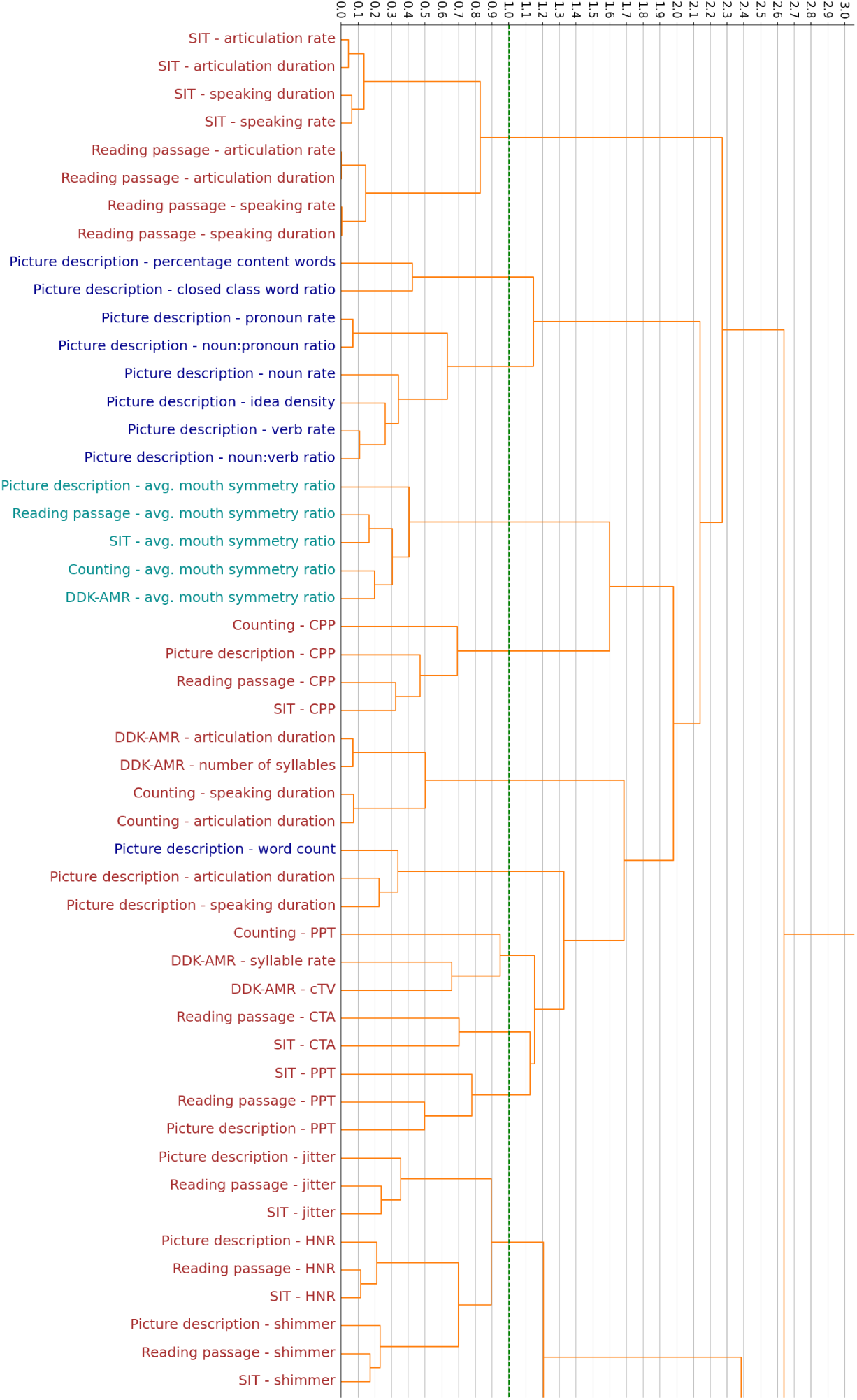
Dendrogram for visualizing feature clusters of acoustic, linguistic, and facial features (exemplary section with 11 out of 27 clusters). The dashed line shows the distance threshold for splitting the clusters.

Next, a representative feature for every cluster was selected to form the final feature set. Receiver operating characteristic (ROC) curve analysis was used in a 5-fold cross validation setup to determine the area under the ROC curve (AUC) for distinguishing bulbar onset participants from non-bulbar onset participants (for every individual feature). 5-fold cross validation was used to ensure generalizability. We implemented it with sklearn’s *Stratified-GroupKFold* function, where the samples were stratified by the class label non-bulbar/bulbar onset, and it was ensured that there is no overlap of a participant’s data between training and test folds. Table 3 shows the clusters and the selected representative features. To further filter features, we imposed a minimum threshold for the ROC-AUC. Only features with an AUC*≥* 0.65 and for which the MCID was larger than the standard error of the mean (SE) were considered in the longitudinal analysis (section 4.6).^10^

**Table 3:**
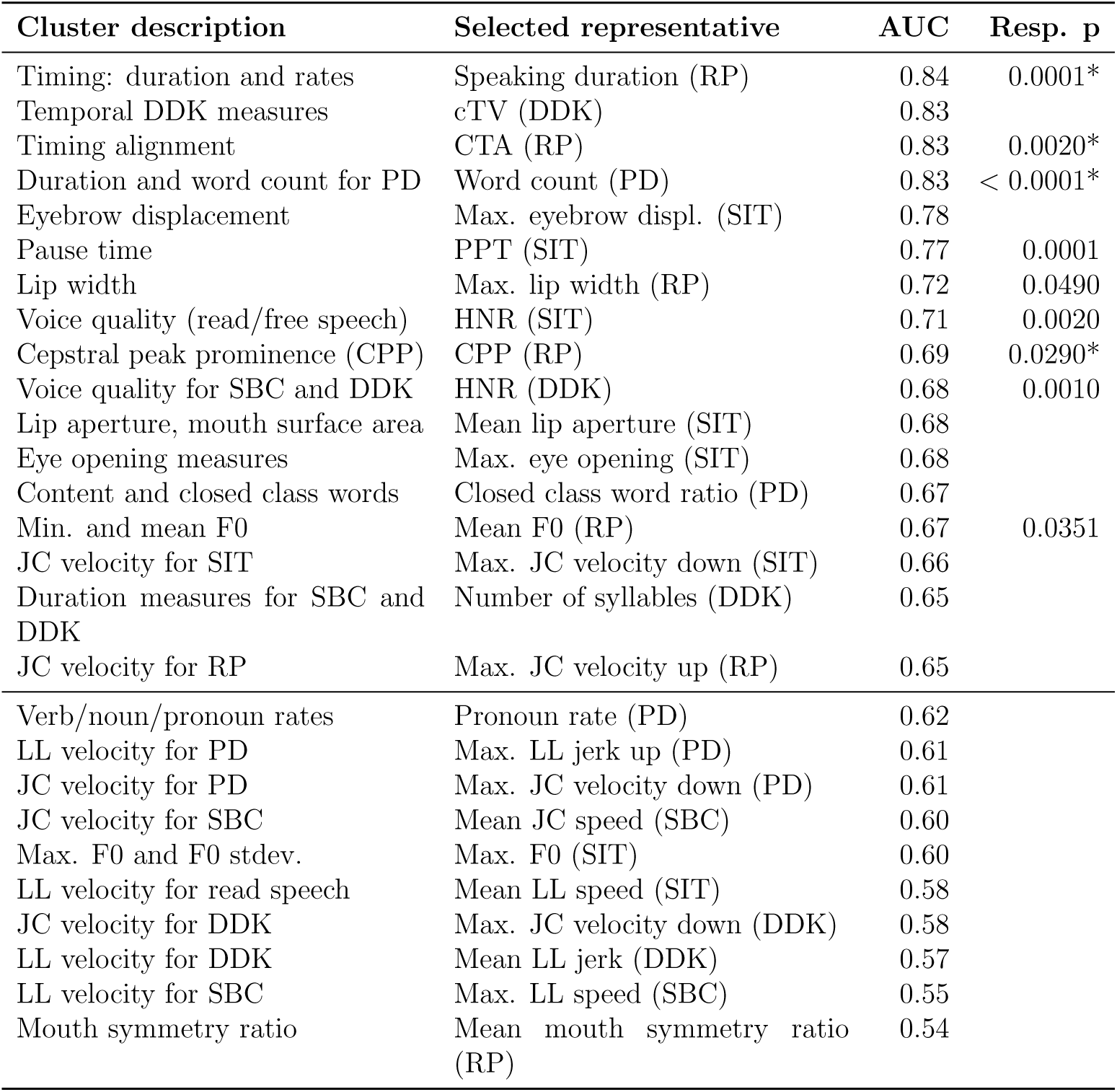
Feature clusters from hierarchical clustering and the selected representative features. *AUC* represents the mean AUC for distinguishing bulbar onset and non-bulbar onset pALS samples across five cross validation folds. Only features with AUC*>* 0.65 were included in the longitudinal analyses. *Resp. p* are the p-values of the responsiveness analysis (see Table 4) and an asterisk (*) indicates features that showed signal in the sensitivity analysis. LL: lower lip, JC: jaw center, RP: reading passage, DDK: diadochokinesis, PD: picture description, SBC: single breath counting, SIT: sentence intelligibility test.

### 4.6. Longitudinal analysis

Responsiveness and sensitivity of features over time was evaluated using growth curve models (GCMs) [70], which provide a linear fit for a non-linear mixed effects model to estimate the trajectory of a metric over time with random slopes and intercepts for each participant [28]. Growth curve models produce estimates of smoothed trajectories of change over time by using observed repeated measures of each individual, making it the ideal statistical method to answer the research questions posed in this paper. The assumption here is that a latent growth process (functional decline) is responsible for the change in observed measures. GCM fitting was performed in R. GCM curves for distinct cohorts can help identify differences in the longitudinal trajectory of measures in the two cohorts. In this study, more than 80% of participants had at least 3 repeated measures, thus minimising any impact of variability in the number of sessions per participant on the growth curve models [71].

#### 4.6.1. Responsiveness

For the responsiveness analysis, the two cohorts chosen for growth curve modeling were sessions from pALS with bulbar onset and those from pALS with non-bulbar onset. First, for every selected feature, we examined whether the rate of change (or slope of the linear fit) is significantly different between the two cohorts. Then, for those features that showed differences, responsiveness was evaluated in two ways: (i) the time taken in weeks to detect deterioration greater than the standard error of the mean for the cohort (statistical utility) and (ii) the time taken in weeks to detect deterioration greater than the MCID value (clinical utility).

#### 4.6.2. Sample Size

To investigate the relationship between responsiveness and sample size of the participant cohort, sample sizes of 30, 25, 20, 15 and 10 participants were randomly sampled 100 times, without replacement, from both cohorts^11^. GCMs were run for each of these 100 iterations. Mean responsiveness for a sample size was calculated by taking the average slope for each cohort across the 100 iterations.

#### 4.6.3. Correlations

To explore the relationship between responsive metrics and the ALSFRSR scale, we ran Spearman correlations between metrics that showed differences in slopes of bulbar and non-bulbar onset pALS and the ALSFRS-R total score, ALSFRS-R bulbar subscore and the ALSFRS-R speech question [5].

#### 4.6.4. Sensitivity

For sensitivity analysis, we wanted to ask whether these metrics can detect bulbar speech deterioration even during those intervals of time where patients report no speech changes on the ALSFRS-R instrument. The two cohorts analysed for this purpose were sessions from healthy controls and all contiguous pALS sessions with a speech score of 3. We decided to look at pALS sessions with a speech score of 3 because these pALS were deemed to exhibit bulbar impairment (albeit per self-perception) but still had speech that was intact enough for objective analysis. A feature was determined to be sensitive if the slope of the GCM for pALS with a steady speech score of 3 varied as compared to the slope of participants from the control cohort with a steady speech score of 4. Note that longitudinal data may be confounded by the presence of learning effects due to the repetition of the same tasks over time. For example, in the case of the Bamboo passage, familiarity with the words in the passage may lead to a decreased speaking duration. The advantage of comparing the trajectory of metrics in ‘clinically-stable’ pALS with that in controls is that it will demonstrate a difference in slopes over any learning effects (assuming the learning effects are equal across cohorts).

## 5. Results

Out of the 17 features selected (after the procedures described in section 4.5), 9 features showed differences in slopes between bulbar onset and non-bulbar onset pALS with the bulbar onset cohort exhibiting a steeper slope (see Figure 4). Details of the slopes per cohort and responsiveness in terms of time to detect change can be found in Table 4. RP speaking duration was found to be the measure with the most responsive statistical utility (2.11 weeks in pALS with bulbar onset). When both statistical and clinical utility are taken into account, RP CTA was the most responsive measure in both cohorts. RP CTA showed statistical and clinical utility in detecting changes in bulbar onset pALS within less than 4 weeks and in non-bulbar onset pALS within 9 weeks. These results are consistent with what was reported in [30] where only timing-related metrics of the RP were considered. PD word count also seemed responsive in detecting changes in bulbar onset pALS within less than 11 weeks and in non-bulbar onset pALS within 9 weeks. However, the shorter duration in the non-bulbar onset cohort is due to an increase in word count over time. This could perhaps be attributed to participants getting familiar with the PD task over repeated sessions and thus exhibiting a learning effect. However, the bulbar onset cohort shows a sharp decrease in PD word count over time despite any learning effect, thus capturing the rapid decline of articulatory and perhaps respiratory function. RP speaking duration demonstrated good responsiveness in detecting statistical changes in both cohorts and clinical change in the bulbar onset cohort. However, when it comes to detecting clinical change in the non-bulbar onset cohort, it takes 23.5 weeks. Although all other features also show differences in the longitudinal trajectory between bulbar onset and non-bulbar onset pALS, the time taken to observe a clinical change, especially in non-bulbar onset pALS, may be too long to be of clinical utility for some interventional trials.

**Figure 4:**
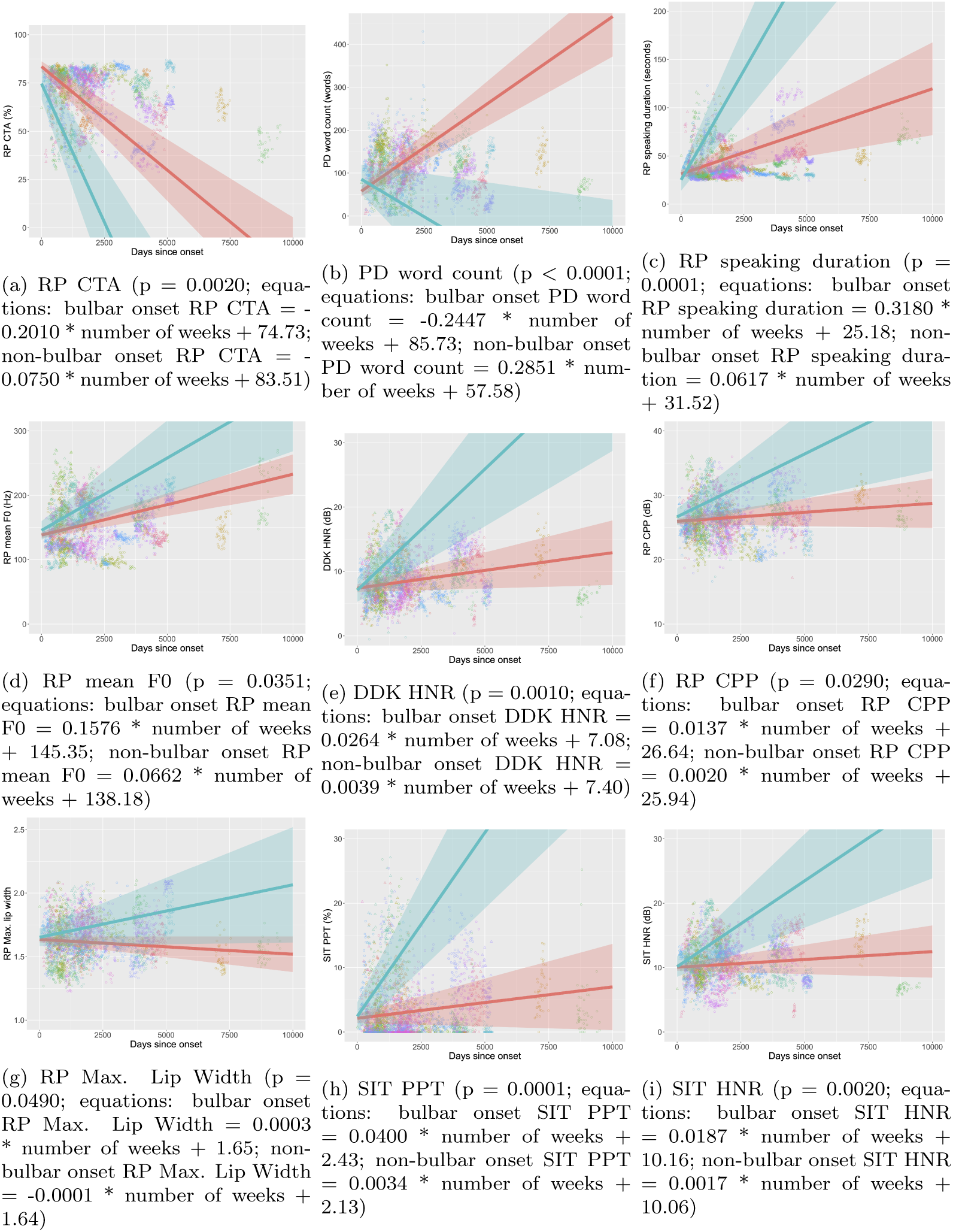
Growth curve models showing rates of change for bulbar onset pALS (blue) as compared to non-bulbar onset pALS (red). Note: The cohort-specific lines in the growth curve model figures are not linear regression fits. They represent the average intercept and slope across all participants in the respective cohorts. Each data point represents a session.

**Table 4:** Responsiveness of metrics. Bulbar onset: n = 36 (598 sessions), Non-Bulbar onset: n = 107 (2790 sessions)

| Metric | MCID | Onset | Slope ± standard error<br>of slope per week | Standard error (SE)<br>of the mean | Weeks to detect<br>change > SE | Weeks to detect<br>change > MCID |
| --- | --- | --- | --- | --- | --- | --- |
| RP CTA<br>(%) | 0.66 | Bulbar | -0.2010 ± 0.0408 | 0.48 | 2.34 | 3.28 |
| | | Non-Bulbar | -0.0750 ± 0.0185 | 0.23 | 3.07 | 8.8 |
| PD word count<br>(words) | 2.5 | Bulbar | -0.2447 ± 0.1296 | 1.56 | 6.38 | 10.22 |
| | | Non-Bulbar | 0.2851 ± 0.0573 | 0.93 | 3.26 | 8.77 |
| RP speaking duration<br>(seconds) | 1.45 | Bulbar | 0.3180 ± 0.0654 | 0.67 | 2.11 | 4.56 |
| | | Non-Bulbar | 0.0617 ± 0.0310 | 0.31 | 5.02 | 23.5 |
| RP mean F0<br>(Hz) | 1.82 | Bulbar | 0.1576 ± 0.0433 | 1.48 | 9.39 | 11.55 |
| | | Non-Bulbar | 0.0662 ± 0.0181 | 0.67 | 10.12 | 27.49 |
| DDK HNR<br>(dB) | 0.44 | Bulbar | 0.0264 ± 0.0068 | 0.15 | 5.68 | 16.67 |
| | | Non-Bulbar | 0.0039 ± 0.0031 | 0.06 | 15.38 | 112.82 |
| RP CPP<br>(dB) | 0.41 | Bulbar | 0.0137 ± 0.0054 | 0.13 | 9.49 | 29.93 |
| | | Non-Bulbar | 0.0020 ± 0.0024 | 0.06 | 30 | 205 |
| RP Max. lip width | 0.01 | Bulbar | 0.0003 ± 0.0002 | 0.0069 | 23 | 33.33 |
| | | Non-Bulbar | -0.0001 ± 0.0001 | 0.0029 | 29 | 100 |
| SIT PPT<br>(%) | 0.96 | Bulbar | 0.0400 ± 0.0093 | 0.23 | 5.75 | 24 |
| | | Non-Bulbar | 0.0034 ± 0.0042 | 0.07 | 20.59 | 282.35 |
| SIT HNR<br>(dB) | 0.59 | Bulbar | 0.0187 ± 0.0055 | 0.16 | 8.56 | 31.55 |
| | | Non-Bulbar | 0.0017 ± 0.0025 | 0.05 | 29.41 | 347.06 |

Mean responsiveness of RP speaking duration, PD word count, RP CTA and RP mean F0 remains stable, with narrow confidence intervals, even with sample sizes as low as 15 per cohort (see Figure 5). However, we observed that the *uncertainty about this estimate* generally increases as the sample size decreases. For all other features, the number of weeks required to detect a statistical and clinical change in the non-bulbar cohort is either unstable or too large to be of any clinical utility. Surprisingly, for some of the metrics, responsiveness was greater at a sample size of 10 than that of 15. We think this could be a result of model overfitting and may not be generalizable.

**Figure 5:**
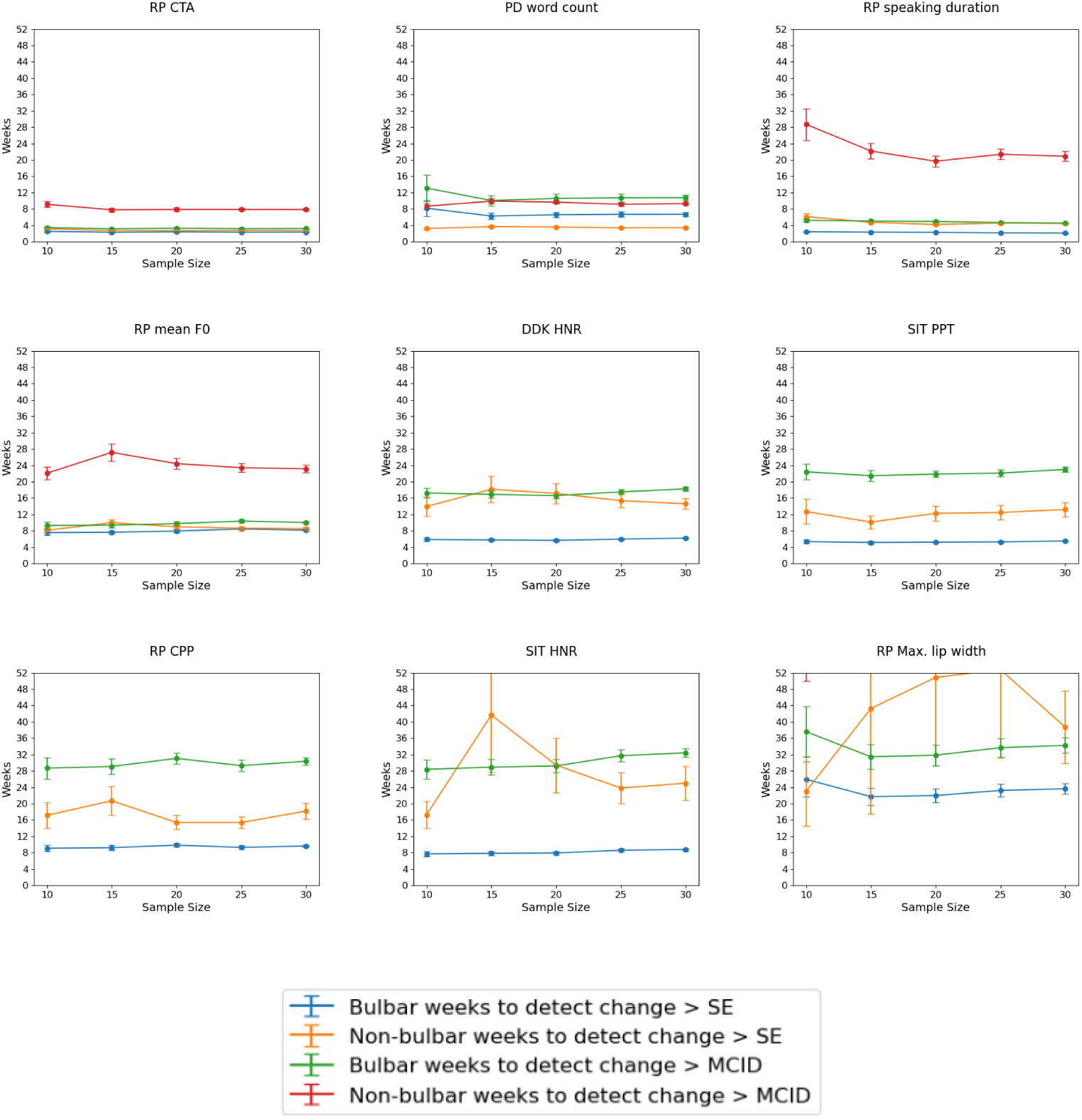
Weeks required to detect a change greater than SE and MCID as a function of sample size. For these plots, the vertical limit of the y-axis was set to 52 weeks. Any metric that requires more than 52 weeks or 1 year to detect statistically or clinically-important changes may not be useful. The red curve is thus missing in many of the subfigures.

Certain speech metrics (like RP speaking duration, RP CTA and SIT PPT) showed moderate to strong correlations with the ALSFRS-R speech question score and the ALSFRS-R bulbar subscore but not the ALSFRS-R total score (see Figure 6).

**Figure 6:**
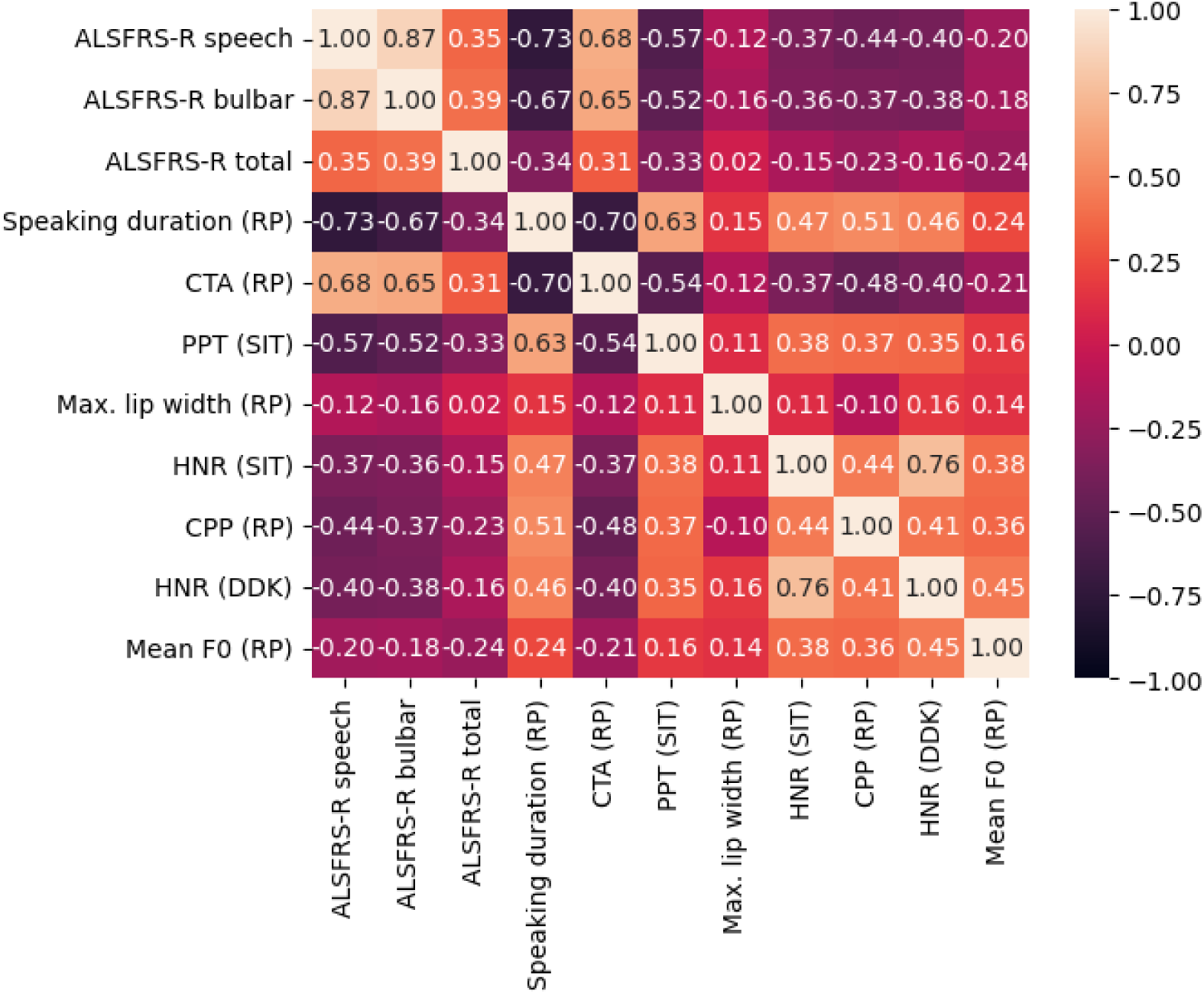
Correlation matrix with Spearman’s *ρ* values indicating the correlation between responsive metrics and ALSFRS-R scores.

Four features were sensitive enough to show a longitudinal change before any change in the ALSFRS-R speech score of patients from 3 (see Table 5) when compared to controls. These features were: RP speaking duration, PD word count, RP CPP and RP CTA. However, for three of these four features (RP speaking duration, PD word count and RP CTA), a learning effect in controls can be observed through the negative slope for RP speaking duration and RP CTA and a positive slope for PD word count. Since clinical deterioration of speech in controls is not expected, any changes in features can be attributed to familiarisation with the task or learning effects. Note that the slope for CTA is negative in controls because an increase in speaking rate would reduce the CTA value because the elicitation will be faster than the canonical elicitation of the reading passage. Differences between controls and pALS were observed despite the presence of these learning effects.

**Table 5:** Sensitivity of metrics. 47 pALS (793 sessions), 135 controls (3428 sessions). Controls had a slope significantly different from 0 for metrics with an asterisk (*) next to them. pALS had a slope significantly different from controls for metrics with two asterisks (**) next to their p-value of difference.

| Metric | p-value of difference | Cohort | Intercept ± standard error | Slope ± standard error |
| --- | --- | --- | --- | --- |
| RP speaking duration*<br>(seconds) | < 0.0001** | Controls<br>pALS | 34.55 ± 0.87<br>44.96 ± 1.88 | -0.0368 ± 0.0101<br>0.1153 ± 0.0220 |
| RP CTA*<br>(%) | 0.0017** | Controls<br>pALS | 79.92 ± 0.85<br>66.81 ± 1.83 | -0.0384 ± 0.0113<br>-0.1130 ± 0.0238 |
| PD word count*<br>(words) | 0.0071** | Controls<br>pALS | 100.63 ± 4.47<br>70.94 ± 9.72 | 0.6204 ± 0.0692<br>0.2355 ± 0.1430 |
| RP CPP<br>(dB) | 0.0341** | Controls<br>pALS | 25.64 ± 0.21<br>27.37 ± 0.47 | -0.0055 ± 0.0029<br>0.0073 ± 0.0060 |
| DDK HNR<br>(dB) | 0.1047 | Controls<br>pALS | 7.11 ± 0.23<br>9.62 ± 0.49 | 0.0055 ± 0.0033<br>0.0171 ± 0.0071 |
| SIT PPT<br>(%) | 0.1584 | Controls<br>pALS | 1.66 ± 0.34<br>4.71 ± 0.73 | -0.0036 ± 0.0040<br>0.0083 ± 0.0084 |
| RP mean F0*<br>(Hz) | 0.1885 | Controls<br>pALS | 147.23 ± 3.09<br>156.85 ± 6.33 | 0.0417 ± 0.0198<br>0.1007 ± 0.0449 |
| SIT HNR<br>(dB) | 0.2657 | Controls<br>pALS | 10.01 ± 0.22<br>12.10 ± 0.47 | -0.0002 ± 0.0025<br>0.0059 ± 0.0055 |
| RP Max. lip width<br>(Hz) | 0.9014 | Controls<br>pALS | 1.65 ± 0.01<br>1.69 ± 0.03 | -0.0001 ± 0.0001<br>-0.0001 ± 0.0003 |

## 6. Discussion and Conclusions

***Summary.*** We can summarize the modeling results and in turn, the novel contributions of this manuscript, by means of the following concise answers to the research questions framed in Section 1:

1. *Optimal Feature Selection*. Through a comprehensive feature selection process that took multicollinearity into consideration, we selected 17 representative features that were able to distinguish bulbar onset pALS from non-bulbar onset pALS with an AUC *≥* 0.65.
2. *Responsiveness to Longitudinal Change*. 9 of these 17 features showed a significantly different change over time in pALS with bulbar onset as compared to pALS with non-bulbar onset.
3. *Time to Detect Change*. Of these metrics, RP CTA and PD word count were the most responsive, in that our fitted growth curve models for these metrics showed the shortest time to detect a change that was statistically and clinically relevant.
4. *Effect of Sample Size*. Responsiveness of metrics remains relatively stable even with small sample sizes. However, the uncertainty about this estimate generally increases as the sample size decreases.
5. *Sensitivity Relative to Clinical Standard*. Four speech features – RP CTA, PD word count, RP speaking duration and RP CPP – also showed a statistically significant change over time even when the clinical gold standard indicated no clinical change in bulbar-impaired pALS. For this, we chose pALS who perceived their speech to be impaired, i.e., a score of 3 on the ALSFRS-R speech question. Healthy controls showed a learning effect over time for three of these four features (a slope statistically different from 0) – RP speaking duration, PD word count, RP CTA – perhaps because they got more familiar with doing the tasks. Under the assumption that pALS and controls exhibit similar learning effect rates, progression in pALS was significantly different as compared to controls, indicating that these metrics are more sensitive than the clinical gold standard ALSFRS-R at detecting speech deterioration. Future work will examine the veracity of this assumption by carefully modelling learning effects across a larger cohort of ALS patients.

## Implications

These results are promising and could have a direct impact on the future of remote digital assessment and remote patient monitoring in both clinical trials and clinical care. The need for objective and responsive outcome measures for improved and accelerated clinical trials is high. Digital biomarkers that can be collected remotely – and therefore more frequently than standard clinical assessments – serve as promising outcome measures for tracking disease progression and changes over time in an automated, objective, and scalable manner. Our findings on sample size suggest that speech-based digital biomarkers show considerable promise in enabling clinical trial designs with small sample sizes. For the most responsive metrics, while the mean responsiveness is stable with decreasing sample size, the uncertainty about the estimate (standard error) increases marginally as the sample size decreases. This has important implications for clinical trial design, where one desires high confidence in chosen biomarker endpoint estimates on the one hand, and as low a participant sample size as possible on the other. This is because the estimated costs of a clinical trial are directly proportional to the number of participants and clinic visits required [72], and therefore the sample size has to be small enough to fit budgetary constraints. However, an underpowered trial may result in a statistically inconclusive outcome and the failure of the clinical trial. Furthermore, in rare neurodegenerative disorders like ALS, accommodating smaller sample sizes is especially important, given the rapidity of disease progression. Remotely-collected digital biomarkers, showing responsiveness at sample sizes of around 15 participants, not only enable smaller sample sizes but also obviate the need for frequent clinical visits. It is possible, however, that the heterogeneity of disease manifestation and progression in pALS may not always enable clinical trials with small sample sizes. Future studies with diverse populations need to be conducted to confirm the findings of this study. Concerning the generalizability of the presented findings to other neurological or mental disorders, it is important to note that the specific set of selected useful features, including the relative utility of different modalities, will likely be different in other disorders (cf. related work in Parkinson disease [73] or mental health disorders like depression [74] and schizophrenia [75]), as well as the sensitivity analysis with respect to a traditional clinical outcome (which is dependent on the disease). However, the methods that were applied in this study can be readily transferred to other diseases.

## Limitations and future work

It is crucial to interpret the presented findings in the context of the study’s assumptions and limitations. Out of the selected set of multimodal metrics, we found that certain metrics were more useful than others. In particular, facial video metrics showed diminished utility relative to speech metrics. Despite previously presented evidence for the utility of these facial metrics in cross-sectional studies, in ALS and other disorders [26, 27, 76, 77, 75, 74], they were not as responsive to longitudinal change as speech metrics in the specific cohorts investigated in this work, suggesting that these metrics are comparatively less robust than speech metrics for characterizing the strongly heterogeneous nature of ALS disease progression [78]. Some features that were selected during feature selection had an good AUC for discriminating bulbar onset from non-bulbar onset pALS, but were not responsive for longitudinal change, e.g., DDK cTV and SIT maximum eyebrow displacement. While eyebrow placement metrics are not typically expected to be informative about ALS disease progression, we hypothesized good clinical utility for the DDK cTV. A related measure of DDK temporal variability, lip movement jitter, was previously shown in [79] to be useful in distinguishing slow and fast progressors of bulbar ALS, and that we did not observe similar findings for DDK cTV was counterintuitive. On examining the fitted GCM more closely, we observed a much higher variance about the cTV slope estimate for the bulbar onset cohort relative to the non-bulbar cohort, which rendered the differences in slopes statistically insignificant. One possible explanation for this observation could be the challenging nature of estimating cTV automatically with sufficient resolution from audio recordings for the bulbar cohort in particular, which may lead to inaccurate estimates of the true cTV. To contexualize how fine a resolution is required, in our dataset, the cTV feature had a range of approximately 30 to 100ms, and therefore requires very low measurement error to capture accurately, which is challenging even with human annotation.

On a more general note about the selected feature set, we are aware of the fact that other types of features, such as deep neural network based representations, can potentially yield improved performance. However, as interpretability is pivotal in clinical applications, this work focused on well studied and interpretable speech, linguistic, and facial kinematic metrics. Alternative black-box data representations like wav2vec [80, 81] and HuBERT [82] were not in the scope of this study, but should be investigated in future work. Another aspect to consider for future work is the potential effect of speaker sex on certain speech metrics. Frequency related metrics (e.g. F0) and voice quality metrics (e.g. cepstral peak prominence) are influenced by sex; thus, it might be beneficial to analyze such metrics separately for female and male participants.

Methodologically, some necessary assumptions were made to compute the MCID as a measure of clinical utility. How to derive the MCID for a given outcome measure is still an open challenge and multiple approaches to estimate clinically-meaningful change have been proposed [33, 66]. We used the ALSFRS-R speech score as external anchor of meaningfulness and applied ROC analysis to derive MCID thresholds for each feature. It is important to note that the AUC values for these analyses were in the range of 0.51 to 0.64. Such values are not surprising and have been seen in prior work looking at dysarthric speech in ALS [34]. Different approaches and alternatives for an external anchor will be carefully and systematically examined in future work. Also, in our investigation of responsiveness and sensitivity, we applied growth curve models under the assumption of linear trajectories. While this was done for simplicity and ease of interpretation and is still useful, we know that ALS disease progression is often nonlinear [78]. Future work will focus on improving modelling methods to better capture trajectories and the variability in different clusters of patients that may share similar disease progression patterns.

Lastly, it is important to take into account the practical considerations and challenges to overcome for widespread adoption of these multimodal digital biomarker technologies in clinical practice and clinical trials. These include robustness in the face of many different conditions that affect signals from different modalities differently, robustness to atypical speech diversity, heterogeneity and comorbidities involved in progression of disease, recording environments and application settings, and generalizability and statistical power of models as promoted by abundant, good-quality training data [11, 38]. Over and above these, one needs to to overcome several practical challenges, including, but not limited to, privacy (protection, access regulation and security of patient data), economic issues (reimbursement, insurance coverage), clinical issues (potential for lowering quality of care, potential for abuse, fragmentation of care), legal issues (licensure laws, liability concerns) and social issues (differential access to technologies based on socioeconomic background) [83, 84, 38]. While some of these are within our control – for example, ensuring that the Modality platform is HIPAA- and GDPR-compliant, and adheres to strict standards of patient privacy^12^, others will require working as a community to address these gaps in order to accelerate progress towards the next generation of precision digital health.

## Conclusion

In conclusion, we found that the longitudinal trajectories of certain digital speech biomarkers are useful in distinguishing between persons with bulbar onset ALS and non-bulbar onset ALS. These trajectories suggest that clinical change associated with bulbar decline could be detected in a matter of a few weeks in pALS. Among the biomarkers investigated, the timing alignment of read speech as compared to a canonical reading of the passage was the most responsive to bulbar decline. This responsiveness holds true even at low sample sizes. Additionally, some biomarkers are sensitive enough to detect a change before any clinical change is detected by the prevalent gold-standard survey instrument, the ALSFRS-R scale. The findings of this study highlight the importance of including multimodal speech biomarkers from remotely-collected data in clinical trials. Their inclusion can facilitate accessible, speedier and cost-effective randomized controlled trials.

## Data Availability

Data used in the present study is not publicly available.

## Acknowledgements

This work was supported by the National Institutes of Health grant R42DC019877. We thank our collaborators at EverythingALS and the Peter Cohen Foundation for participant recruitment and data collection. We also thank Gabriela M. Stegmann for providing the template R code to fit growth curve models.

## Author contributions

Michael Neumann: Conceptualization; Data curation; Formal analysis; Investigation; Methodology; Validation; Visualization; Writing; Editing.

Hardik Kothare: Conceptualization; Data curation; Formal analysis; Investigation; Methodology; Validation; Visualization; Writing; Editing.

Vikram Ramanarayanan: Conceptualization; Funding acquisition; Formal analysis; Investigation; Methodology; Project administration; Writing; Editing; Supervision.

## Footnotes

1 A note on the terminology used in this paper: We use the term *metric* to denote the general concept of speech and facial characteristics (e.g., speaking rate), and we use the term *feature* to denote a metric that was extracted for a specific stimulus or task (e.g., speaking rate for a reading passage task).

2 https://versiticlinicaltrials.org/salusirb [retrieved on 2023-12-29], protocol number: 2020-06-PI42

3 https://www.everythingals.org/research [retrieved on 2023-12-29]

4 For some participants, there was no match in the healthy controls cohort. As a result, the final dataset contained less healthy controls than people with ALS.

5 Participants filled out the ROADS questionnaire [44] for a subset of sessions instead of the ALSFRS-R. Therefore, the ALSFRS-R score was not available for all sessions.

6 https://spacy.io/ [retrieved on 2023-12-29]

7 https://github.com/rpy2/rpy2 [retrieved on 2023-12-29]

8 https://aws.amazon.com/transcribe/ [retrieved on 2023-12-29]

9 The maximum distance at which all features would be combined into one single cluster was 5.4

10 AUC values were rounded to 2 digits after the decimal point before determining the best feature in each cluster. Among the 17 resulting clusters with AUC*≥* 0.65 there was only one instance of a tie: the features SIT speaking rate, RP speaking duration, and RP speaking rate all had an AUC of 0.84. We ran the responsiveness analysis for all three features and report results for the best performing one, RP speaking duration.

11 We did not consider sample sizes of 40 or greater per cohort for this analysis to avoid sampling the 36 participants in the bulbar cohort more than once.

12 https://www.modality.ai/regulatory/privacy

